# Early COVID-19 Interventions Failed to Replicate 1918 St. Louis vs. Philadelphia Outcomes in the United States

**DOI:** 10.1101/2020.07.02.20145367

**Authors:** Aliea M. Jalali, Brent M. Peterson, Thushara Galbadage

**Affiliations:** Department of Kinesiology and Health Science, Biola University, La Mirada, CA, United States; College of Nursing and Health Care Professions, Grand Canyon University, Phoenix, AZ, United States

**Keywords:** Coronavirus, Spread, Intervention, Prevention, Outcomes, Evidence-based practice, Health Disparities, Influenza virus

## Abstract

The Coronavirus disease 2019 (COVID-19) pandemic has elicited an abrupt pause in the United States in multiple sectors of commerce and social activity. As the US faces this health crisis, the magnitude, and rigor of their initial public health response was unprecedented. As a response, the entire nation shutdown at the state-level for the duration of approximately one to three months. These public health interventions, however, were not arbitrarily decided, but rather, implemented as a result of evidence-based practices. These practices were a result of lessons learned during the 1918 influenza pandemic and the city-level non-pharmaceutical interventions (NPIs) taken across the US. During the 1918 pandemic, two model cities, St. Louis, MO, and Philadelphia, PA, carried out two different approaches to address the spreading disease, which resulted in two distinctly different outcomes. Our group has evaluated the state-level public health response adopted by states across the US, with a focus on New York, California, Florida, and Texas, and compared the effectiveness of reducing the spread of COVID-19. Our assessments show that while the states mentioned above benefited from the implementations of early preventative measures, they inadequately replicated the desired outcomes observed in St. Louis during the 1918 crisis. Our study indicates that there are other factors, including health disparities that may influence the effectiveness of public health interventions applied. Identifying more specific health determinants may help implement targeted interventions aimed at preventing the spread of COVID-19 and improving health equity.

## INTRODUCTION

As the first wave of Coronavirus Disease 2019 (COVID-19) pandemic began to sweep through the United States (US) in March 2020, multiple public health measures were enforced across the nation in an unprecedented manner. However, by the end of June 2020, the US remained one of the largest COVID-19 epicenters, globally, with more than 2.5 million confirmed cases and the number of new daily cases reaching highs in certain states and the US (CDC, 2020b). Now, faced with the renewed threat of experiencing prolonged second wave, many states are reintroducing partial shutdown measures, which are examples of non-pharmaceutical interventions (NPIs). During the first wave of this pandemic, the US strictly implemented multiple NPIs to help mitigate the spread of the disease, and reduce the number of COVID-19-related deaths. Herein we discuss the successes and failures of the implemented evidence-based public health practices amid a nationwide public health crisis that abruptly brought the nation and its economy to a screeching halt.

As of February 2020, while China, Italy, and Spain experienced the turmoil of being the epicenters for the COVID-19 pandemic, the US had only about 50 confirmed cases, and the national populace was nearly unaffected. No one could have anticipated how life was about to change in the ensuing months. In March 2020, different states started to sound the alarms, and place their respective constituencies under states of emergency. After that, increasingly rigorous preventative measures that affected the function and dynamics of societal interaction were implemented. These interventions, aimed at facilitating social distancing and preventing the spread of COVID-19, can be categorized into four broad measures (Galbadage et al., 2020b; Wilder-Smith and Freedman, 2020). These are (1) screening and testing, (2) prevention of mass gatherings, (3) stay at home orders, and (4) the use of face masks. In the US, 44 states of the 50 states implemented statewide stay at home orders at the early stages of the COVID-19 pandemic, paralleling other measures listed above (Figure 1, Supplemental Table 1). The mean duration of stay at home orders for all US states was 49.5 days (SD ± 16.5) (median 50 days, range 25 to 81 days).

**Figure 1.**
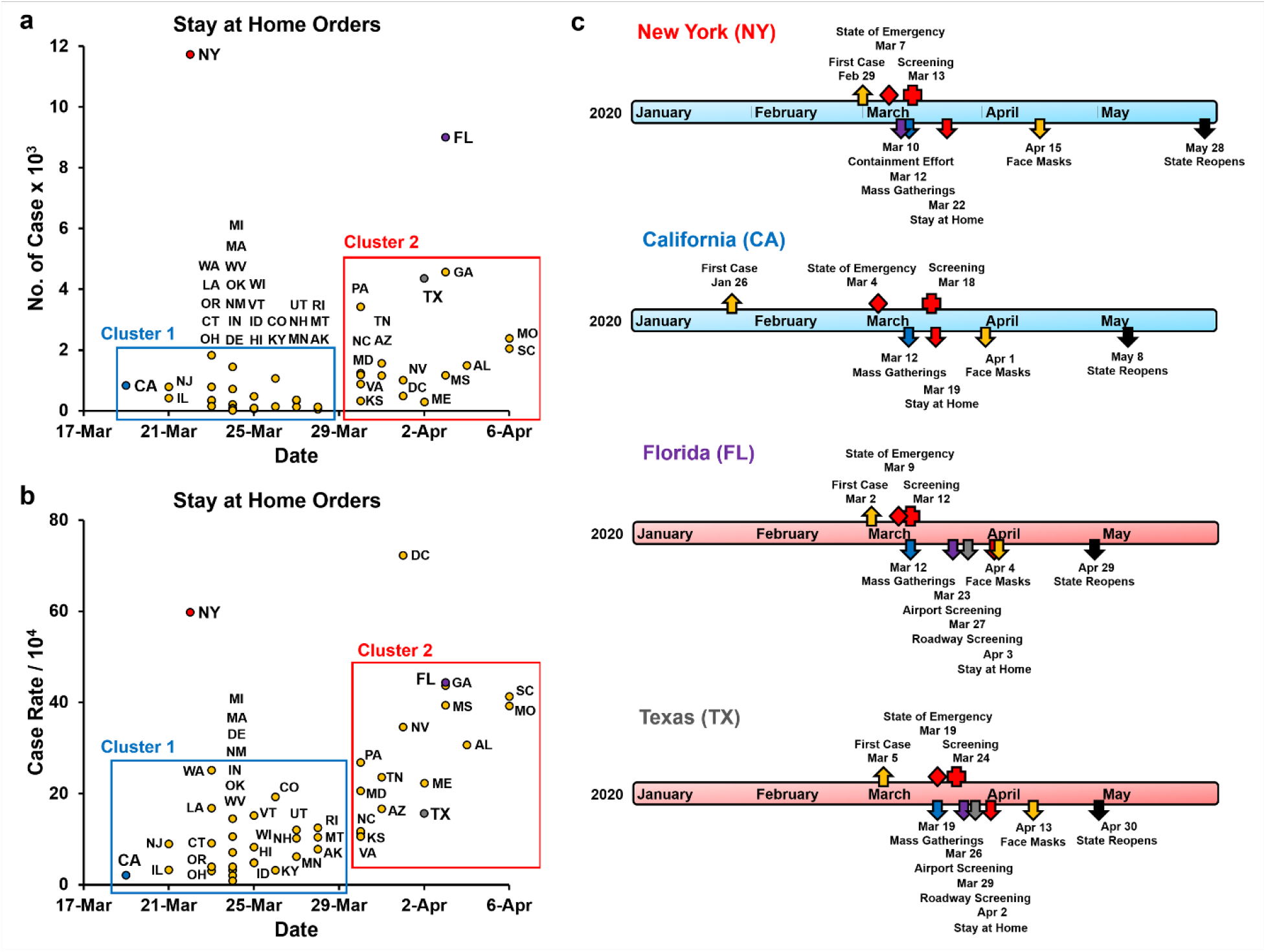
State and county-level public health interventions to contain the spread of COVID-19. (a) The number of lab-confirmed COVID-19 cases at the start of the stay at home orders implemented by each state (Dong et al., 2020). Arkansas, Iowa, Nebraska, North Dakota, South Dakota, and Wyoming did not issue statewide stay at home orders and are not included. Cluster 1 -states that implemented stay at home orders before March 29^th^, 2020, and Cluster 2 - states that implemented these orders after March 29^th^. (b) Case rates of lab-confirmed COVID-19 patients at the start of the stay at home orders implemented by each state. Cases rates are the number of cases per 10,000 of the county population. (c) Timeline of public health response (non-pharmaceutical interventions) in the states of New York (NY), California (CA), Florida (FL) and Texas (TX). These interventions included screening and testing, a ban on mass gatherings, stay at home orders, requirements for face masks in public locations, and other state-specific measures. In NY contained a one-mile containment effort around hotspot New Rochelle in Westchester County. In FL airport and roadway, screening was implemented for travelers coming to FL from the tri-state region as well as other regions with a high prevalence of COVID-19. In TX Airport and roadway, screening was implemented mainly for travelers coming into TX from the tri-state area and Louisiana, where the prevalence of COVID-19 was high. TX did not enforce mandatory use of cloth facemasks at the state level. Travis (4/13), Harris (4/13), Bexar (4/16), Dallas County (4/18) ordered mandatory facemasks.

While seemingly sudden and societally intrusive, historical precedent and evidence-based practices have guided these measures. For example, a century ago, the world experienced a devastating toll on lives caused by the 1918 influenza pandemic. In response to this pandemic, health officials implemented a broad range of NPIs according to the then available understanding of disease transmission (Mills et al., 2004; Ferguson et al., 2005; Markel et al., 2006). Furthermore, studies comparing public health measures implemented by several cities across the United States and other nations such as England further illustrated how these measures helped reduce the spread of the 1918 influenza pandemic and decrease mortality rates (Ferguson et al., 2006; Bootsma and Ferguson, 2007; Handel et al., 2007; Hatchett et al., 2007).

Studies on the 1918 influenza pandemic have focused on contrasting NPIs implemented by two US cities, St. Louis, MO, and Philadelphia, PA. St. Louis imposed strict preventative interventions early on, while Philadelphia minimally applied restrictions at a much later date. Accordingly, St. Louis had a milder outbreak, whereas Philadelphia experienced significantly higher mortality rates (Hatchett et al., 2007). These outcomes observed in the 1918 influenza pandemic helped guide the widely-adopted rigorous public health measures against COVID-19. Hatchett et al. (2007) also identified four critical factors that helped determine the success of the control of the pandemic dissemination. These factors were (1) implementation of early and rapid interventions, (2) duration of the responses, (3) multiple concurrent interventions, and (4) the intensity of the interventions implemented.

Other studies supported these conclusions while emphasizing the effectiveness of early interventions, but also noted that stringent preventative measures could leave many more susceptible individuals once these NPIs are relaxed (Kalnins, 2006; Bootsma and Ferguson, 2007). During the 1918 pandemic, most of the US cities maintained preventative measures for about two to eight weeks (Hatchett et al., 2007). However, cities that relaxed NPIs earlier experienced increased case numbers resulting in second wave resurgences. An inverse relationship between the intensity of the first and second waves of the pandemic was also observed. These observations were partly due to the smaller proportion of susceptible populations present in cities after a strong first wave of the disease (Bootsma and Ferguson, 2007; Hatchett et al., 2007).

Here we compare and contrast public health interventions implemented in the US during the first wave of the COVID-19 pandemic, focusing on four states: New York, Florida, Texas, and California. These states included most of the populous US counties and were affected sharply by the early stages of the COVID-19 pandemic. In addition, we studied the case rates of COVID-19 before, during, and after these measures were implemented, and then compared it to the outcomes of St. Louis, and Philadelphia, during the 1918 influenza pandemic (Figure 2). While variation in the timing and the intensity of the public health measures applied was observed, all four states implemented very similar interventions. Our comparisons show that the early evidence-based interventions implemented by the US were not adequately able to replicate the desired outcomes of St. Louis vs. Philadelphia and curtail the COVID-19 pandemic.

**Figure 2.**
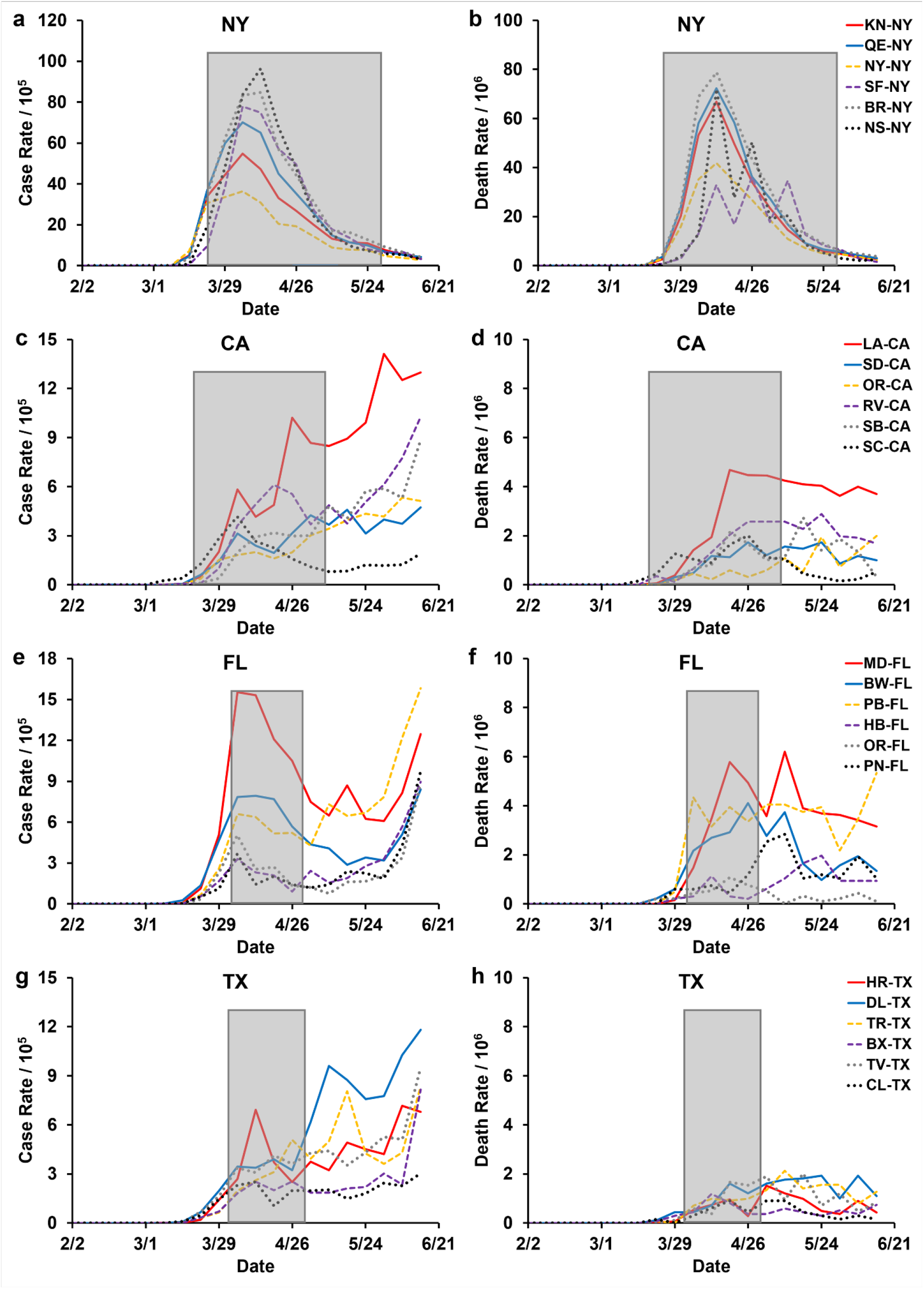
United States COVID-19 cases and mortality in the six most populous counties in the states of New York, California, Florida, and Texas. COVID-19 Cases and deaths are presented as seven-day averages from data provided by Johns Hopkins University and the City of New York (Dong et al., 2020). Grey boxed areas are the duration statewide stay-at-home orders that were implemented by each state: New York (NY) March 22nd to May 28th (68 days), California (CA) March 19th to May 7th (50 days), Florida (FL) April 3rd to April 29th (27 days), and Texas (TX) April 2nd to April 20th (29 days). (a, c, e, g) Case rates are new confirmed COVID-19 cases per 100,000 population in the respective counties. (b, d, f, h) Death rates are new COVID-19 related deaths per 1,000,000 population in the individual counties. (a, b) Six most populous counties in the state of NY: KN-NY - Kings, QE-NY - Queens, NY-NY - New York, SF-NY - Suffolk, BR-NY - Bronx, and NS-NY - Nassau. (c, d) Six most populous counties in the state of CA: LA-CA - Los Angeles, SD-CA - San Diego, OR-CA - Orange, RV-CA - Riverside, SB-CA - San Bernardino, and SC-CA - Santa Clara. (e, f) Six most populous counties in the state of FL: MD-FL - Miami-Dade, BW-FL - Broward, PB-FL - Palm Beach, HB-FL - Hillsborough, OR-FL - Orange, and PN-FL - Pinellas. (g, h) Six most populous counties in the state of TX: HR-TX - Harris, DL-TX - Dallas, TR-TX - Tarrant, BX-TX - Bexar, TV-TX - Travis, and CL-TX - Collin.

## METHODS

### Evaluating the State and County Level Public Health Response

We documented public health responses and actions taken between January 1st and June 30th at the state- and county-level were found using the state and county public health department and government websites. More specifically, we examined press releases and executive orders issued by the state, and the counties were used to determine the public health response. This information collected was organized in an Excel worksheet by date and type of response. We used four broad categories to distinguish between the types of responses. They were (1) screening and testing, (2) prevention of mass gatherings, (3) stay at home orders, and (4) the use of face masks. To have only the most relevant and quantifiable data, we distilled public health responses to critical interventions. Besides, the first positive cases of COVID-19 in each county were also noted. Office Timeline Online was used to create timelines to illustrate this information visually.

### Determining the Case Rates and Mortality Rates

COVID-19 Cases and deaths are presented as seven-day averages from data provided by Johns Hopkins University and the City of New York (Dong et al., 2020). Case rates were calculated as new confirmed COVID-19 cases per 100,000 population in the respective counties. Death rates were calculated as new COVID-19 related deaths per 1,000,000 people in the individual counties. We calculated the case rates and mortality rates in the six most populous counties in the states of New York (Kings, Queens, New York, Suffolk, Bronx, and Nassau), California (Los Angeles, San Diego, Orange, Riverside, San Bernardino, and Santa Clara), Florida (Miami-Dade, Broward, Palm Beach, Hillsborough, Orange, and Pinellas, and Texas (Harris, Dallas, Tarrant, Bexar, Travis, and Collin). We then overlapped the duration statewide stay-at-home orders that were implemented by each state (Grey boxes) to evaluate the ensuing case and mortality rates.

### Public Health Response to COVID-19

As mentioned earlier, responses to earlier pandemics in the US included school closures, restaurant restrictions, emergency declarations, gathering restrictions, stay at home orders, and non-essential business closures (Gupta et al., 2020). The COVID-19-related responses have been mainly relegated to state-level decision making and based on necessity and intensity within each state. To characterize the state-level COVID-19 interventions, we compared and contrasted the broad measured implemented by the states of California, Florida, New York, and Texas.

#### Screening and Testing

Targeted screening for COVID-19 began in California and New York with Los Angeles (LAX), San Francisco (SFO), and New York (JFK) airports for travelers coming from Wuhan, China, starting on January 17^th^ (CDC, 2020e). The first reported case in the US occurred on January 26^th^ in California. New York, Florida, and Texas all had initial cases within the first week of March (Figure 1c). Early in the pandemic, testing was limited, and priority was given to high-risk individuals, including symptomatic patients, healthcare workers, first responders, essential workers, and individuals in contact with other high-risk individuals. As more tests were readily available, fewer restrictions were placed on who was able to get tested (Florida Department of Health, 2020; State of California, 2020b; State of New York, 2020a; Texas Department of State Health Services, 2020). In addition to walk-up and drive-through sites, mobile testing sites were also deployed in Florida and New York to increase the number of tests administered (City of New York, 2020; Florida Division of Emergency Management, 2020). Each state also implemented contact tracing to identify potentially exposed individuals (CDC, 2020c).

#### Mass Gatherings

The next primary public health intervention implemented across all four states was the cancellation of mass gatherings of 250 individuals, followed by 50 individuals per location (Supplemental Tables 2-5). These orders followed shortly after initial cases were identified in each state. Events that brought in large amounts of attendance, such as concerts, sporting events, and festivals were canceled first. Next, the states incrementally decreased the number of people allowed to gather in one location until, eventually, the state recommended that people should only interact with those who were within the same household.

#### Stay at Home Orders

One of the most rigorous measures utilized during COVID-19 was the stay at home orders. California was under stay at home order for 50 days (March 19th to May 7^th^) (State of California, 2020a). The stay at home order in California was implemented more rigorously at the county level because the state-level order acted more as a recommendation (Supplemental Table 3). The NY “State on PAUSE” plan stay at home order was enforced for 68 days (March 22nd to May 28^th^) before the state started its Phase one reopening plan (Cuomo, 2020; State of New York, 2020b; c). Florida state stay at home order was in effect for 27 days (April 3rd to April 29^th^) (State of Florida, 2020). Texas implemented stay at home orders for 29 days before relaxing these measures statewide (April 2nd to April 30^th^) (Abbott, 2020).

Many US states enacted stay at home orders very early on in the COVID-19 transmission. States with early COVID-19 cases placed these measures before April 29^th^ (cluster 1) and did so with a statewide case count of fewer than 2000 cases, while states that put stay at home orders after April 29^th^ did so before reaching 5000 cases (cluster 2) (Figure 1a, Supplemental Table 1). When adjusted to the county population, these measures were implemented with case rates of below 50 cases per 10,000 (Figure 1b). The only exception was New York, which implemented these measures after 11,700 cases were confirmed. (Figure 1a).

#### Cloth Face Masks

On April 3^rd^, the Centers for Disease Control and Prevention (CDC) released its recommendation for all individuals to use cloth face masks when in public (CDC, 2020a). The goal of this recommendation was to reduce the viral transmission from asymptomatic carriers that may unknowingly spread to disease to susceptible individuals (Esposito et al., 2020; Galbadage et al., 2020b). While the extent to which the effectiveness of this measure is debatable, it helps bring more awareness to the public and help curtail the person-to-person transmission of the virus (Eikenberry et al., 2020). California was the first to implement this statewide on April 1^st^, which was two days before the CDC’s recommendation (Figure 1c). New York also implemented this measure as a state-level order, but it happened two weeks after the CDC’s recommendation. Florida and Texas only recommended face coverings at the state-level but was mandated in most counties (supplemental Tables 4 and 5).

### Differences in Statewide Responses to COVID-19

The public health interventions implemented across the four states, New York, California, Florida, and Texas, were very similar. Any differences stem from the relative time of implementation and the intensity of measures taken. Unfortunately, New York was one of the first states severely affected by COVID-19 and was likely too late to implement these preventative measures (Figure 1a and b, Figure 2 and b). The initial wave of COVID-19 in New York, therefore, resembled that of Philadelphia during the 1918 pandemic. California, on the other hand, initiated precautionary measures early and seemed to follow the outcomes of St. Louis, at least in the initial stages (Figure 2, c, and d). Regulations in both of these states were more stringent, and often had consequences such as fines and jail time tied to not adhering to them.

In Texas and Florida, the implementation of specific public health interventions was less rigorous as compared to California and New York. In Texas, for example, the regulations were not implemented as quickly or as firmly at the state-level. Some public health interventions, such as the ban on gathering, stay at home orders and wearing cloth face masks, may have been perceived as violations of individual liberties and disrupting businesses. In many ways, the small-government philosophy of these states left essential decisions and actions to be made at the county-level. Around the time many states went into shut down mode, spring break activities remained open in Florida. The decision to not shut down before spring break was made in support of the state’s economy. It was only after large tourist attractions, including Universal Studios and Disney World, decided to close were more rigorous measures put in place in Florida.

### The Spread of COVID-19 Across States and Counties

During the first months of COVID-19, the disease spread rapidly across the United States. In New York, the number of positive cases grew exponentially over the first month of the pandemic, especially in the New York City area and surrounding boroughs. However, unlike other states, the number of daily cases in New York has decreased consistently since the end of April. In California, Florida, and Texas, the number of daily cases has continued to increase over time at a slower rate compared to New York. To better understand the dynamics of COVID-19 spread in each of these states, we reviewed the number of cases and deaths in the six most populous counties in each of these states (Figure 2).

In New York, the most populous counties all experienced a similar first wave of COVID-19, with a peak of about 100 cases per 10,000 people in early April (Figure a). Most counties in the state of California continued to have a relatively slow, but steady rise in the number of cases, making it difficult to distinguish between a first and a second wave (Figure 2c). We observed a similar pattern in the counties in Florida and Texas, except Miami-Dade County in Florida, which showed a peak case rate of about 15 cases per 10,000 people in early April (Figure 2c, e, g). Among these states, it is clear that New York experienced a robust first wave and a negligible second wave of the COVID-19 pandemic. While California, Florida, and Texas were spared from a significant first wave with cases rate peaking at less than 20 cases per 10,000, they are now facing a much higher risk for a prolonged second wave of the disease.

### US COVID-19 Interventions Failed to Replicate 1918 Pandemic Outcomes

In the COVID-19 pandemic, the goal of effective public health preventative measures implemented was to mitigate and contain the spread of the disease. In the US, for the most part, public health interventions followed the principles of effective NPIs. They were implemented early on in the pandemic, using multiple preventative measures, with high intensity and for average durations longer than 45 days (Figure 1, Supplement Table 1). The exception to this was New York, which delayed the initiation of these measures (Figure 1a and b). This caused New York to experience a peak first wave, with hospitals reaching their capacity and a peak number of deaths occurring during mid-April (Figure 1b). However, New York enforced its preventative measures for close to three months, which in turn helped them bring their daily case rates to less than 5 cases per 10,000 by the end of June.

In contrast to New York, most other states followed the evidence-based recommendations, as stated above (Figure 1). This helped states “flatten the curve” to various degrees and control the initial spread of COVID-19 within their states. However, these public health interventions seemed to have only prolonged the transmission potential of the COVID-19 as states, including California, Florida, and Texas were experiencing new daily highs in confirmed cases by the end of June 2020 (CDC, 2020b). While the general expectation was that US states would follow the outcome of St. Louis during the 1918 pandemic, they have fallen short of replicating this desired outcome. On the contrary, by the end of June 2020, many such states were reimplementing statewide partial shutdown measures to prevent a potential second wave of COVID-19.

## DISCUSSION

While the United States failed to prevent the early spread of COVID-19 effectively, some countries had better success containing the Coronavirus with their public health interventions. In Iceland, for example, when cases were identified, public health officials implemented the following strategies: quarantine requirements for international travelers, rigorous tracing of infection, ban on gatherings larger than 20 persons, school closures with limited openings of elementary and preschools, defining areas of higher risk, and regular communication with the general public (Iceland Directorate of Health, 2020). New Zealand, another island nation with great success, was more rigorous in the process by modifying and intensifying pre-existing plans for the management of influenza pandemics from previous outbreaks (Baker et al., 2020). These methods included the declaration of a national emergency, a nationwide lockdown, closure of non-essential work locations, banning social gatherings, extreme restrictions on travel, and closure of all schools. Furthermore, as part of this intensified strategy, border security was also tightly regulated. However, there are distinct differences between Iceland, New Zealand, and the United States. Iceland and New Zealand are small island nations with much smaller populations, making it much easier to implement rigorous preventative measures, including better travel restrictions and contact tracing. They were also able to coordinate their public health response more consistently nationwide, unlike the US, which enforced COVID-19 interventions mainly at the state level.

Several factors can help explain why the US was unable to effectively replicate the outcomes of St. Louis vs. Philadelphia during the 1918 flu pandemic. These include (1) the level of adherence to these implemented preventative measures and social behaviors, (2) disparities in social determinants of health, and (3) extensive global and domestic travel with little restrictions. Regardless of the public health intervention intensity, they can be ineffective if people do not consistently adhere to them. Besides, numerous risk factors have been identified for COVID-19 and its clinical outcomes. These include advanced age, sex, immune-compromised status, and comorbidities, including chronic respiratory diseases, diabetes, and hypertension (Galbadage et al., 2020a; Li et al., 2020; Richardson et al., 2020). American Indians, African Americans, and Hispanic individuals have been reported to be four to five times more likely to be hospitalized for COVID-19 when compared to non-Hispanic whites (CDC, 2020d). Disparities in social determinants of health, such as access to healthcare, uninsured population, employment, poverty, education, and population density, can also contribute to the differences observed in COVID-19 transmission. Potential clusters of these risk factors and health determinates present in different geographic regions can lead to the disproportionate spread of the Coronavirus. In conclusion, it is crucial to consider factors such as adherence to preventative measures, and health disparities, in evaluating the effectiveness of COVID-19 interventions implemented. These factors likely caused the US early COVID-19 public health measures to be less effective in containing the Coronavirus pandemic and is an important further direction of research.

## Data Availability

All datasets presented in this study are included in the article/ supplementary material.

## Author Contributions

AJ analyzed the US public health response and helped with the figure preparation, writing, and editing of the manuscript. BP aided in intervention comparative analysis and helped with the writing and editing of the manuscript. TG aided in disease spread, and comparative analysis, and helped with the figure preparation, writing and editing of the manuscript.

## Acknowledgments

We thank Drs. Richard S. Gunasekera, Genti Buzi, and Biola research students JongWon See, Alexis Gulsvig, and Sumaia Khoury for their discussions and data analysis on this research topic. We also thank Dr. Jeffrey S. Wang, Infectious Disease Specialist at Kaiser Permanente, Anaheim, California, for his clinical insights.

## Supplementary Material

The Supplementary Material for this article can be found below.

